# Cortical Similarities in Psychiatric and Mood Disorders Identified in Federated VBM Analysis via COINSTAC

**DOI:** 10.1101/2023.09.28.23296219

**Authors:** Kelly Rootes-Murdy, Sandeep Panta, Ross Kelly, Javier Romero, Yann Quidé, Murray J. Cairns, Carmel Loughland, Vaughan J. Carr, Stanley V. Catts, Assen Jablensky, Melissa J. Green, Frans Henskens, Dylan Kiltschewskij, Patricia T. Michie, Bryan Mowry, Christos Pantelis, Paul E. Rasser, William R. Reay, Ulrich Schall, Rodney J. Scott, Oliver J. Watkeys, Murray J. Cairns, Gloria Roberts, Philip B. Mitchell, Janice M. Fullerton, Bronwyn J. Overs, Masataka Kikuchi, Ryota Hashimoto, Junya Matsumoto, Masaki Fukunaga, Perminder S. Sachdev, Henry Brodaty, Wei Wen, Jiyang Jiang, Negar Fani, Timothy D. Ely, Adriana Lorio, Jennifer S. Stevens, Kerry Ressler, Tanja Jovanovic, Sanne J. H. van Rooij, Sergey Plis, Anand Sarwate, Vince D. Calhoun

## Abstract

Psychiatric disorders such as schizophrenia, major depressive disorder, and bipolar disorder have areas of significant overlap across multiple domains including genetics, neurochemistry, symptom profiles, and regional gray matter alterations. Various structural neuroimaging studies have identified a combination of shared and disorder-specific patterns of gray matter (GM) deficits across these different disorders, though few direct comparisons have been conducted. Given the overlap in symptom presentations and GM alterations, these disorders may have a common etiology or neuroanatomical basis that may relate to a certain vulnerability for mental illness. Pooling large data or heterogeneous data can ensure representation of several participant factors, providing more accurate results. However, ensuring large enough datasets at any one site can be cumbersome, costly, and may take many years of data collection. Large scale collaborative research is already facilitated by current data repositories and neuroinformatics consortia such as the Enhacing NeuroImaging Genetics through Meta-Analysis (ENIGMA) consortium, institutionally supported databases, and data archives. However, these data sharing methodologies can still suffer from significant barriers. Federated approaches can augment these approaches and mitigate some of these barriers by enabling access or more sophisticated, shareable and scaled up analyses of large-scale data which may not be shareable and can easily be scaled up with the number of sites. In the current study, we examined GM alterations using Collaborative Informatics and Neuroimaging Suite Toolkit for Anonymous Computation (COINSTAC). Briefly, COINSTAC (https://coinstac.trendscenter.org) is an open-source decentralized analysis application that provides a venue for analyses of neuroimaging datasets without sharing individual level data, while maintaining granular control of privacy. Through federated analysis, we examined T1-weighted images (N = 3,287) from eight psychiatric diagnostic groups across seven sites. We identified significant overlap in the GM patterns of individuals with schizophrenia, major depressive disorder, and autism spectrum disorder. These results show cortical and subcortical regions, specifically the bilateral insula, that may indicate a possible shared vulnerability to psychiatric disorders.

## Introduction

Mental illness can be severe, negatively impact cognitive and social functioning, and may relate to years of suffering at the helm of misdiagnosis, medication trials, and evolving clinical presentations. Understanding the etiology and the related cortical underpinnings of these disorders can direct advances in treatment. However, another challenge is psychotic disorders and mood disorders can have similar presentations and can overlap across multiple domains including symptoms, genetic predispositions, and regional cortical alterations (American Psychiatric Association, 2013). A recent review of the DSM-5 found that the clinical symptoms that repeated the most across diagnoses were (in order of frequency) insomnia, difficulty concentrating, hypersomnia, psychomotor agitation, and depressed mood (Forbes et al., 2023). Schizophrenia (SZ), affective disorders, autism spectrum disorder (ASD), and post-traumatic stress disorder (PTSD) all vary in presentations but share the categorization of cognitive impairments, varying affective symptoms, and behavioral dysfunction.

Structural neuroimaging studies have separately identified unique cortical alterations associated with certain psychiatric disorders largely noting patterns of cortical deficits. Individuals with SZ have consistent regions of gray matter (GM) deficits in the bilateral insula, anterior temporal lobe, and the medial frontal lobe (Gupta et al., 2015) whereas individuals with bipolar disorder (BP) have shown GM deficits in the medial superior frontal gyrus and gyrus rectus but more GM in the cortico-striato-cerebellar and default mode networks (DMN) (Long et al., 2022). Psychiatric neuroimaging has also identified regions of similarities between some psychiatric disorders, indicating a possible shared cortical underpinning. Even between SZ and BP, similar brain correlates were identified in the bilateral insula, cingulate gyrus, cerebellum, thalamus, vermis, and the supplemental motor cortices (Lee et al., 2020; Rootes-Murdy, Edmond, et al., 2022). A voxel-based morphometry (VBM) study found more gray matter concentration in the putamen of individuals with diagnoses of PTSD, unipolar depression, psychosis, and obsessive-compulsive disorder and that this finding was correlated with symptom severity (Gong et al., 2019). Common patterns of GM deficits in the dorsal anterior cingulate and bilateral insula were identified in a meta-analysis of voxel-based studies comparing SZ, BP, unipolar depression, addiction disorders, obsessive-compulsive disorder, and anxiety disorders (Goodkind et al., 2015). Given the overlap in symptom presentations and specifically, the similarities in gray matter alterations, some of these disorders may have a common neuroanatomical basis that may branch into different diagnostic categories. In this study, we sought to examine the unique and similar cortical alterations across eight different psychiatric and developmental diagnoses (described below) using a federated analysis approach.

### Psychiatric Disorders and their related Symptoms

SZ is characterized by cognitive, behavioral, and emotional dysfunction (American Psychiatric Association, 2013; Andreasen & Flaum, 1991). Symptom presentations include varying combinations of positive symptoms (hallucinations, delusions), negative symptoms (anhedonia, apathy, low mood), cognitive dysfunction (difficulties with abstract thinking, memory deficits, disorganized thinking), and abnormal motor behavior (agitation or catatonia). Structural neuroimaging studies found regional gray matter reductions throughout the cortex (Gupta et al., 2015; Honea et al., 2005; Rootes-Murdy et al., 2021; van Erp et al., 2018) with the largest GM effects identified in the superior temporal gyrus (Gupta et al., 2015) and the cerebellum (Moberget et al., 2018) in individuals with SZ.

BP is a mood disorder marked by extreme polar mood states, from depression to hypomania to mania (American Psychiatric Association, 2013). BP has different classifications defined mainly by severity of mood states; for example, Type I indicates fluctuation from depressed to manic states and Type II indicates fluctuations from depressed to hypomanic states (American Psychiatric Association, 2013). Both BP and major depressive disorder (MDD) can present with impaired cognitive abilities, appetite changes, and psychosis (Kennedy, 2008). Structural neuroimaging shows similar volumetric reductions across the cortex in BP to those identified in SZ with smaller effect sizes (Bora, 2015; de Zwarte et al., 2019; Murray et al., 2004; Rootes-Murdy, Edmond, et al., 2022).

Major depressive disorder (MDD) is another prevalent mood disorder associated with negative symptoms of anhedonia, apathy, and low mood (American Psychiatric Association, 2013). MDD is the leading cause of disease burden worldwide and has a lifetime prevalence of 5 – 17% (2022). Structural studies show inconsistent findings in MDD in the hippocampus and amygdala, with a caveat of smaller volumes largely being attributed to age more than the disorder. However, advanced brain aging was observed across the cortex in MDD (Han et al., 2021; Jaworska et al., 2014, 2016; Schmaal et al., 2016).

PTSD is categorized under the stress-disorder group in the DSM-5 and can develop after a person has experienced one or more traumatic events (American Psychiatric Association, 2013). PTSD symptoms typically presents along four clusters: re-experiencing or trauma memory reliving, avoidance, trauma memory reliving, persistent negative emotions, cognitive biases, and sleep disturbances largely related to hyperarousal (American Psychiatric Association, 2013). The prevalence rate of PTSD in the United States is 3.6% with female presenting with higher rates (5.2%) then males (1.8%) (Goldstein et al., 2016). The trauma association with PTSD has resulted in numerous imaging studies focusing on regions of interest (ROI) related to fear/threat circuitry, particularly the amygdala and hippocampus, where volumes are largely reduced when compared to healthy controls (Kunimatsu et al., 2020; Logue et al., 2018; Shin, 2006). In addition to the fear circuit, significant effects of PTSD have also been identified in the insula, anterior cingulate cortex, superior frontal gyrus, and the temporal gyri (Kunimatsu et al., 2020).

Autism Spectrum disorder (ASD) is a development disorder that lies along a spectrum of deficits in social communication, repetitive behaviors, and cognitive deficits (American Psychiatric Association, 2013). The current prevalence of ASD in the US is estimated at one in every 59 individuals (Baio et al., 2018). Structural MRI studies have identified increases in the cortical thickness of the frontal lobe and reduced cerebellar volume in individuals with ASD. However, there are also inconsistent findings reported across the cortex, including both increases and decreases in the hippocampus, amygdala, basal ganglia, and thalamus (Pagnozzi et al., 2018).

Mild cognitive impairment (MCI) is defined as cognitive decline that is greater than expected (given age and education) but does not significantly impact or interfere with functioning (American Psychiatric Association, 2013). Current estimates of prevalence range from 3% to 19% in adults older than 65 years (Farlow, 2009; Gauthier et al., 2006). MCI is considered a risk state for dementia because more than half of individuals diagnosed with MCI progress to some form of dementia within 5 years. There is limited research focused solely on MCI, but structural studies have identified nodes specific to MCI (as opposed to Alzheimer’s disease) within the parietal lobe, cerebellum, and the occipital lobe (Hojjati et al., 2019).

### Comorbidity Across Diagnoses

Although these disorders are classified as separated diagnoses, there are many symptoms, genetic, and neuroanatomical features that are shared by multiple disorders. Previous studies have even examined the Diagnostic Statistical Manual (DSM) to explore the possible comorbidity among diagnoses (Borsboom et al., 2011; Tio et al., 2016). The symptoms of sleep disturbance, difficulty concentrating, psychomotor agitation, and depressed mood overlap across several psychiatric disorders including but not limited to SZ, MDD, PTSD, and BP (Forbes et al., 2023). There is also symptomatic overlap between SZ and ASD; both disorders can present with deficits in social cognition and have a male sex bias (Santos et al., 2022; Sugranyes et al., 2011).

There are also common genetic and structural features that have been identified between SZ and BP (Cross-Disorder Group of the Psychiatric Genomics Consortium et al., 2013; Mahon et al., 2012; Padmanabhan et al., 2015; Potash et al., 2003; Rootes-Murdy, Edmond, et al., 2022). Structural neuroimaging studies have identified similarities in the regions of gray matter (GM) deficits in individuals with SZ and BP (<u>Doan et al., 2017</u>; <u>Schwarz et al., 2019</u>; <u>Sorella et al., 2019</u>; <u>Lee D.-K. et al., 2020</u>; <u>Cheon et al., 2022</u>). These two disorders may be better described along a continuum of varying cognitive deficits and psychosis (<u>Jabben et al., 2010</u>; <u>Hill et al., 2013</u>). The genetic variations shared between SZ and BD is 15%, between BD and MDD is 10%, and between SZ and MDD is 9% (Cross-Disorder Group of the Psychiatric Genomics Consortium et al., 2013). In addition to similarities, there are often comorbid presentations across disorders. MDD is the most commonly seen in individuals with post-traumatic stress disorder (PTSD) when compared to all other psychiatric disorders (Rytwinski et al., 2013). Although, it should be noted that there is some evidence that depressive and anxiety disorders may increase the risk for PTSD and vice versa (Spinhoven et al., 2014). Therefore, the comorbidity may not be the presentation of independent disorders but more a shared etiology, shared set of risk factors, or even symptom overlap. Future research is needed to examine the shared and unique cortical alterations of these disorders.

### Unaffected Relatives

Unaffected relatives of individuals with psychosis have shown inconsistencies in cognitive tasks with some relatives having similar impairments to those affected and some performing similar to healthy controls (Bora & Pantelis, 2013; Montag et al., 2012). Structural studies identified the left orbitofrontal cortex (OFC) and right cerebellum as regions in both BP and relatives of BP that were significantly smaller than healthy controls (Eker et al., 2014). In fMRI studies, striatal dysfunction was related to poorer performance in individuals with SZ however, in the unaffected relatives this same dysfunction was thought to relate to a compensatory pattern because it was not related to poor performance (Vink et al., 2006). Given these presentations, unaffected relatives may offer insight into both a heritable risk load but also, correlates of resistant factors associated with psychiatric disorders.

### Federated approaches

Single studies tend to have small effects, masking true results, or even leading to false results. Open data and consortia like ENIGMA have helped move beyond these limitations, but there are many datasets (often highly relevant clinical data sets) which are unable to be openly shared (or shared at all), and high-dimensional (e.g., voxelwise analysis) or iterative approaches (e.g., classification/machine learning) that are challenging or impossible to perform using a manual consortia approach. Federated analysis as in the Collaborative Informatics and Neuroimaging Suite Toolkit for Anonymous Computation (COINSTAC) tool mitigates this issue by allowing analysis of data that would be otherwise inaccessible, hence increasing the sample size, without moving data from its original location or exposing the individual subject data to the other consortia members (Rootes-Murdy, Gazula, et al., 2022).

The aim of this project was to leverage federated analyses to examine transdiagnostic presentations in individuals with either schizophrenia, bipolar disorder, major depressive disorder, post-traumatic stress disorder, mild cognitive impairment, autism spectrum disorder, or unaffected relatives of individuals with psychosis with the aim of uncovering possible relatedness and distinct mechanisms underneath their respective diagnoses.

## Methods

We performed a large-scale multisite meta-analysis of subcortical gray matter concentration alterations in seven psychiatric and developmental disorders and unaffected relatives compared to healthy controls using COINSTAC (http://coinstac.trendscenter.org).

### Participants

This study included data from 3,287 individuals; 572 individuals with schizophrenia (SZ), 189 individuals with mild cognitive impairment (MCI), 147 unaffected relatives of an individual diagnosed with psychosis, 121 individuals with bipolar disorder (BP), 96 individuals classified as spectrum (further information about this category can be found in the Supplemental Material), 88 individuals with autism spectrum disorder (ASD), 44 individuals diagnosed with major depressive disorder (MDD), 32 individuals with post-traumatic stress disorder (PTSD), and 1,998 healthy controls (HC) from the following datasets, many previously described in the literature; the Australian Schizophrenia Research Bank (ASRB; Loughland et al., 2010), Emory University Grady Trauma Project (Emory; Fani et al., 2012, 2019; Stevens et al., 2013), Imaging Genetics in Psychosis (IGP; Quidé et al., 2022), Centre for Healthy Brain Aging-Sydney Memory and Aging Study (MAS; Sachdev et al., 2010), Older Australian Twins Study (OATS; Sachdev et al., 2009) Cognitive Genetics Collaborative Research Organization (COCORO consortium; (Koshiyama et al., 2020), and Bipolar Kids and Sibs-Sydney (Sydney; (Roberts et al., 2013). Diagnoses were confirmed by clinical physicians at each respective site as part of the study’s protocol. All data were collected under approval of local institutional review boards and all participants provided informed consent. The original study designs are described in previous publications (cited above). See Table 1 for more details on participant information including age, sex, and diagnostic breakdown from each site and see the Supplemental Materials section for the participant inclusion and exclusion criteria for each site.

**Table 1.** Participant Information Across Sites.

| **SITE** | **N** | **AGE (SD)** | **SEX (M%)** | **DIAGNOSES (N)** | **HC** |
| --- | --- | --- | --- | --- | --- |
| **ASRB** | 321 | 39.36 (11.72) | 204 (63.55%) | 215 SZ; 14 Spectrum | 92 |
| **COCORO** | 1719 | 33.89 (14.45) | 891 (51.83%) | 305 SZ; 88 ASD; 82 Spectrum; 44 MDD | 1200 |
| **EMORY** | 105 | 38.23 (11.30) | 0 | 32 PTSD | 73 |
| **IGP** | 179 | 37.94 (11.85) | 88 (49.16%) | 64 BP; 52 SZ | 63 |
| **MAS** | 449 | 78.28 (4.69) | 198 (44.10%) | 164 MCI | 285 |
| **OATS** | 193 | 70.33 (5.17) | 71 (36.79%) | 25 MCI | 168 |
| **SYDNEY** | 321 | 21.64 (5.05) | 134 (41.75%) | 147 Unaffected Relatives; 57 BP | 117 |
| **TOTAL** | 3287 | 41.79 (10.84) | 1586 (48.25%) | 572 SZ; 189 MCI; 147 Unaffected Relatives; 121 BP; 96 Spectrum; 88 ASD; 44 MDD; 32 PTSD | 1998 |

SD: Standard deviation; M: male; HC: healthy controls; SZ: schizophrenia; ASD: Autism Spectrum disorder; MDD: major depressive disorder; PTSD: post-traumatic stress disorder; BP: bipolar disorder; MCI: mild cognitive impairment

### MRI Dataset Acquisition and Preprocessing

T1-weighted scans were obtained at the seven sites with varying MRI field strength and vendors. Details regarding MRI acquisition and sample demographics can be found in Table 2. Individual sites obtained approval from local institutional review boards and informed consent was obtained at the time of the original study. All preprocessing steps were completed within the COINSTAC (http://coinstac.trendscenter.org; Ming et al., 2017; Plis et al., 2016) federated analysis tool (see below for description). All T1-weighted images used the following preprocessing protocol for data harmonization. Images were co-registered and normalized to the standard Montreal Neurological Institute (MNI) template using a 12-parameter affine model, resliced to a voxel size of 2 mm X_J2 mm X 2 mm and segmented into gray matter, white matter, and cerebrospinal fluid using Statistical Parametric Mapping 12 (SPM12; http://www.fil.ion.ucl.ac.uk/spm/software/spm12/). All images were smoothed at 10 mm FWHM prior to analyses. The preprocessing resulted in a total of 3,287 gray matter concentration images (Meda et al., 2008; Segall et al., 2009).

**Table 2.**
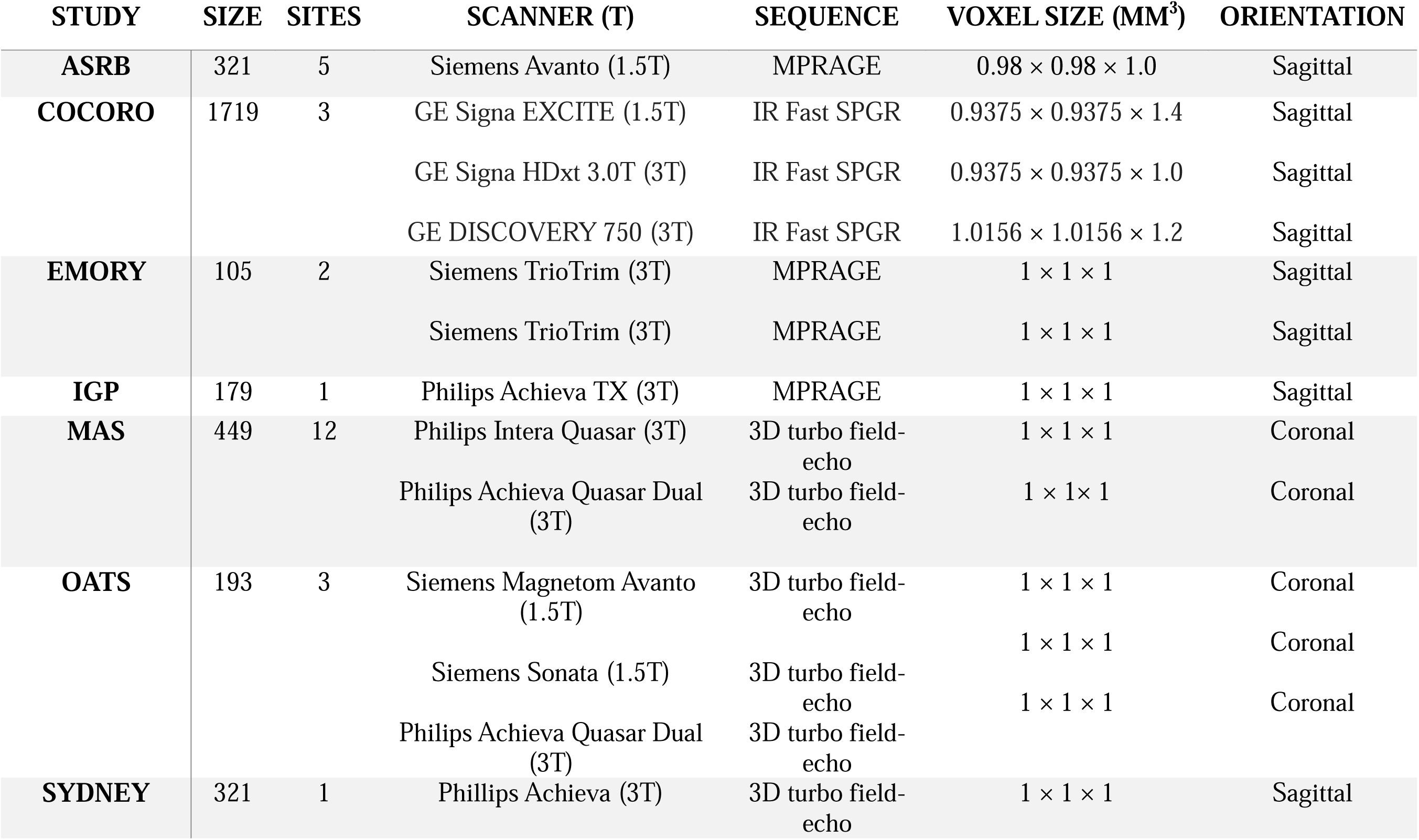
MRI Scanner Information Across Sites.

### COINSTAC

In the current study, we examined the gray matter alterations using the COINSTAC tool (Ming et al., 2017; Plis et al., 2016; Rootes-Murdy, Gazula, et al., 2022). Briefly, COINSTAC (https://coinstac.trendscenter.org) is an open-source decentralized analysis application that provides a venue for analyses of neuroimaging datasets without sharing any individual level data, while maintaining granular control of privacy. COINSTAC has been utilized for multiple decentralized approaches circumventing the need for pooling data. Instructions for installing the application and running analyses can be found online at https://github.com/trendscenter/coinstac-instructions. See Figure 1 for a breakdown of the COINSTAC architecture.

**Figure 1.**
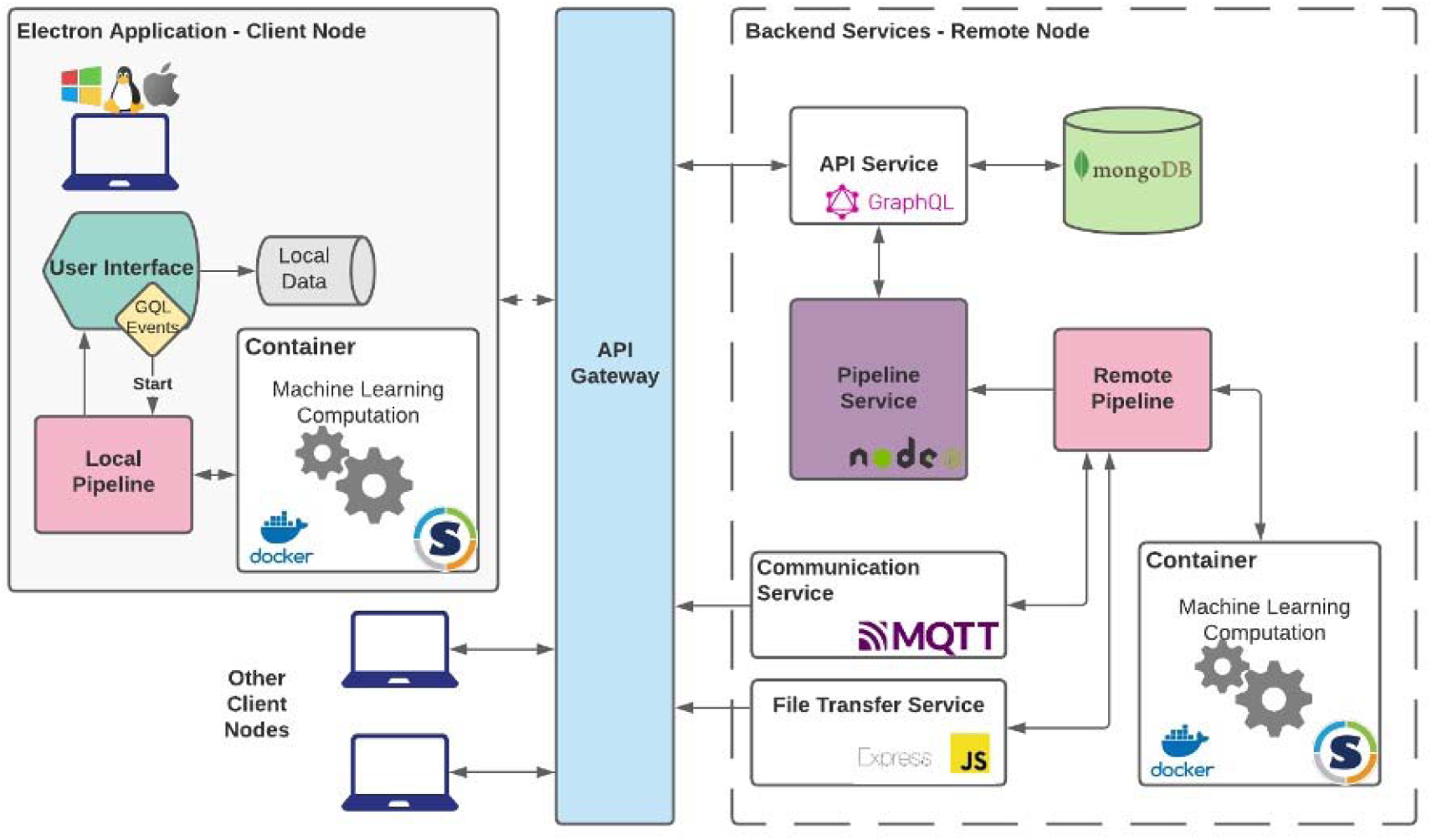
The COINSTAC architecture. Figure 1 shows a diagram of the COINSTAC architecture, including both client nodes and remote services. The COINSTAC application uses standard web communication protocols to transfer (meta)data between each client node and a publicly accessible remote node via an API gateway. Client nodes and the remote node run computations in a pipeline, with client nodes sending metadata derived from local data and the remote node aggregating and running required computations, sending results back to client nodes for either final output or iteration if required. Pipeline results are stored in a noSQL data store provided by MongoDB.

In this study, seven global sites contributed T1-weighted images (N = 3,287) for a voxel-based morphometry (VBM) analysis within the COINSTAC platform. Data harmonization was completed using the COINSTAC software with all the sites using the same preprocessing pipeline and then all scanning sites were added as covariates to the regression models. The regression models were also run in COINSTAC allowing sites to perform all computations on their local data and intermediate results are collected by a cloud-based “private aggregator” which synthesized the results and returned them to the sites. The −log(*p*) maps of each diagnostic gray matter image were compared to one another using spatial correlations to examine cross-diagnostic similarities and differences.

### General Linear Regressions

We employed a general linear model on all normalized and smoothed gray matter images to associate each diagnosis with voxelwise gray matter concentration, controlling for age, sex, site, and apolipoprotein-E (APOE) allele classification. Each diagnostic category was compared separately resulting in eight separate regression models. Following the regressions, the −log(*p*) maps of each diagnostic gray matter image were compared using spatial correlations.

## Results

In general, comparing across the diagnoses, there was common tendency of gray matter concentration reductions in the disorder groups except for the unaffected relatives. The gray matter reductions were largely seen in the bilateral insula, medial prefrontal cortex, parahippocampal gyrus, and rectus. Below is a breakdown of the results for each diagnostic category. All results listed below are compared to healthy controls.

### SZ Results

Individuals with SZ (N = 572) had global reductions across the entire cortex when compared to healthy controls and had the largest effect size of any diagnosis. The results showed reduced gray matter concentration in the bilateral insula, cingulate, parahippocampal gyrus, and the ventromedial prefrontal cortex compared to healthy controls. See Figure 2 for more details. *BP Results*

**Figure 2.**
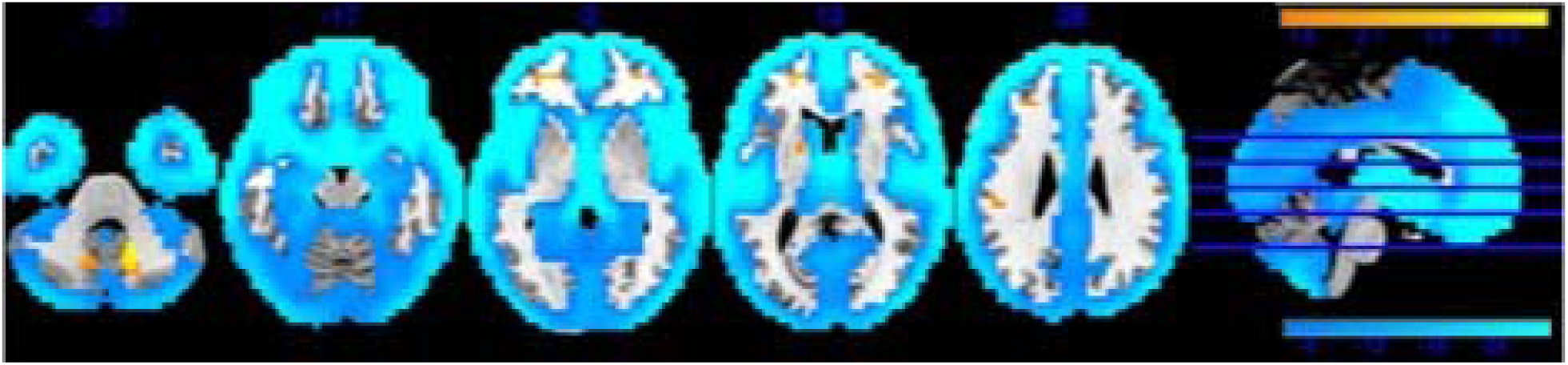
Gray Matter Alteration in Schizophrenia

Individuals with BP (N = 121) showed GM concentration reductions centered around the bilateral insula and ventromedial prefrontal cortex, these reductions were similar to those identified in SZ, but to a smaller effect. There was also a small cluster of GM reductions in the occipital lobe. There were no areas of increased GM concentration in these individuals compared to healthy controls. See Figure 3 for more details.

**Figure 3.**
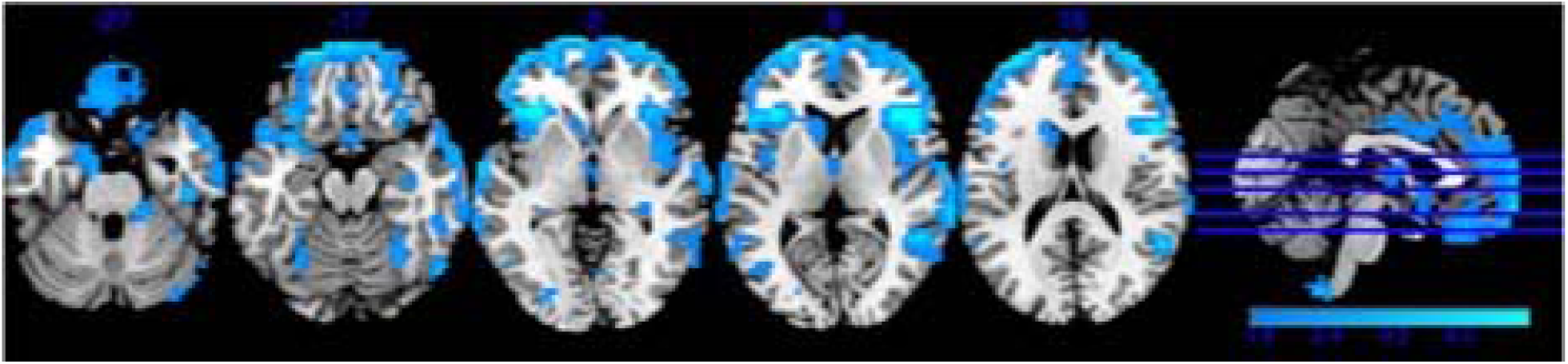
Gray Matter Alteration in Bipolar Disorder

### MDD Results

Individuals with major depressive disorder (N = 44) had clusters of reduced gray matter concentration in the bilateral insula, cerebellum, and the cingulum, rectus, medial prefrontal cortex (peak of cluster: x = 1, y = 55, z = −6). There were no clusters of increased GM concentration when compared to healthy controls. See Figure 4 for more details.

**Figure 4.**
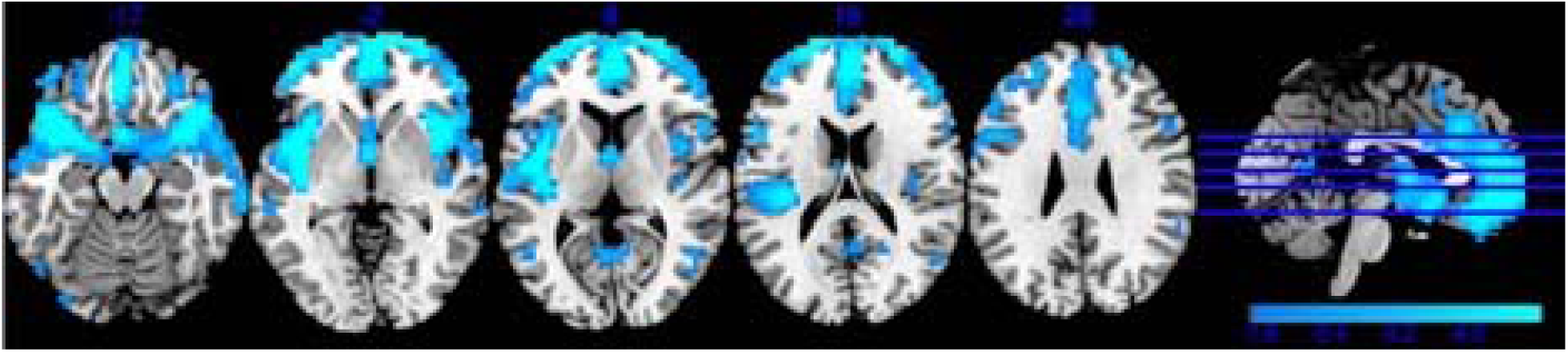
Gray Matter Alteration in Major Depressive Disorder

### PTSD Results

Individuals with PTSD (N = 32) had less gray matter concentration in the cingulate, precuneus, left insula, retrosplenial cortex, and the bilateral superior occipital gyrus. There were clusters of increased GM identified in the bilateral superior parietal lobules (Peak of R cluster: x = 25, y = −55, z = 62). See Figure 5 for more details.

**Figure 5.**
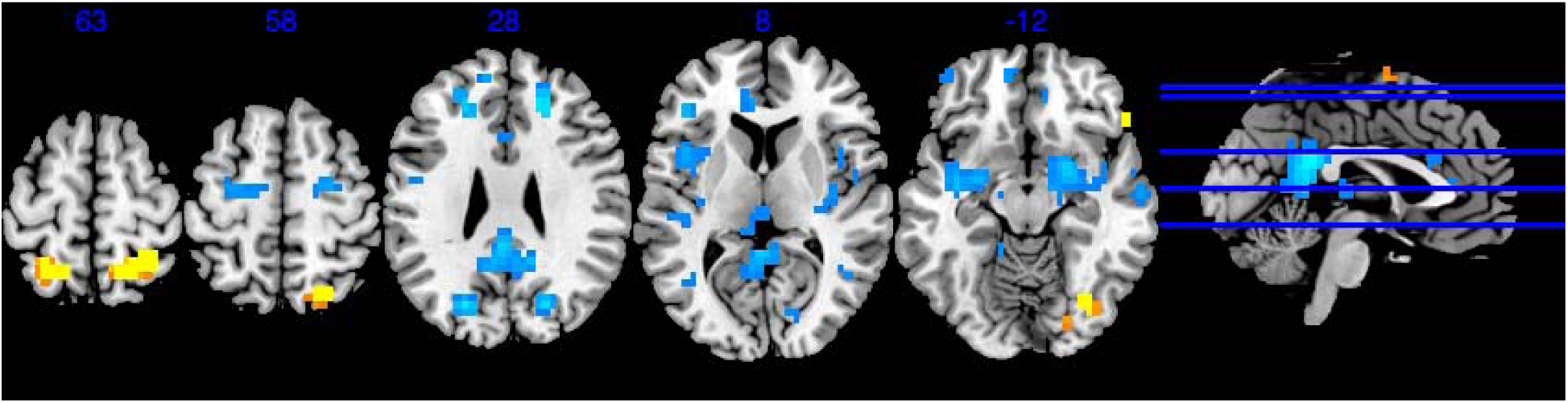
Gray Matter Alteration in Post-Traumatic Stress Disorder

### ASD Results

Individuals with ASD (N = 88) had small clusters of gray matter reductions in the bilateral insula, parahippocampal gyrus, and Heschl’s gyrus. There were also reductions identified in the olfactory and fusiform gyri. Individuals with ASD showed a small cluster of GM increase in the right putamen and the cerebellum IX. See Figure 6 for more details.

**Figure 6.**
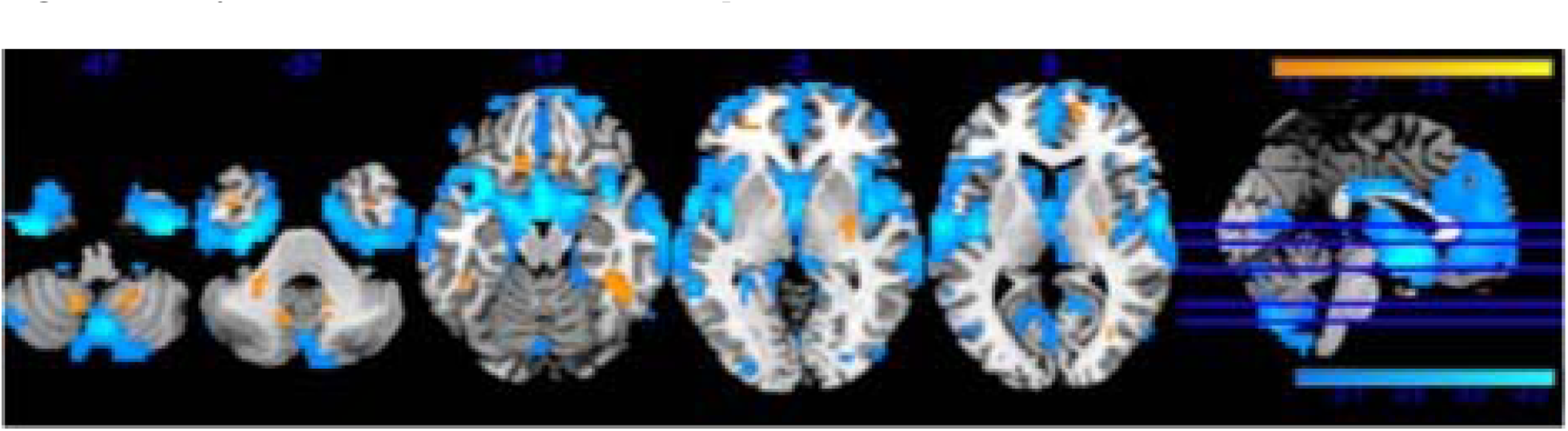
Gray Matter Alteration in Autism Spectrum Disorder

### MCI Results

Individuals with MCI (N = 189) also had global reductions in subcortical regions, but with a smaller effect size than individuals with SZ. Results showed reduced gray matter in the bilateral hippocampus, cerebellum VI, Crus I, amygdala, cingulum, and the bilateral insula.

There were no notable increases in GM concentration. See Figure 7 for more details.

**Figure 7.**
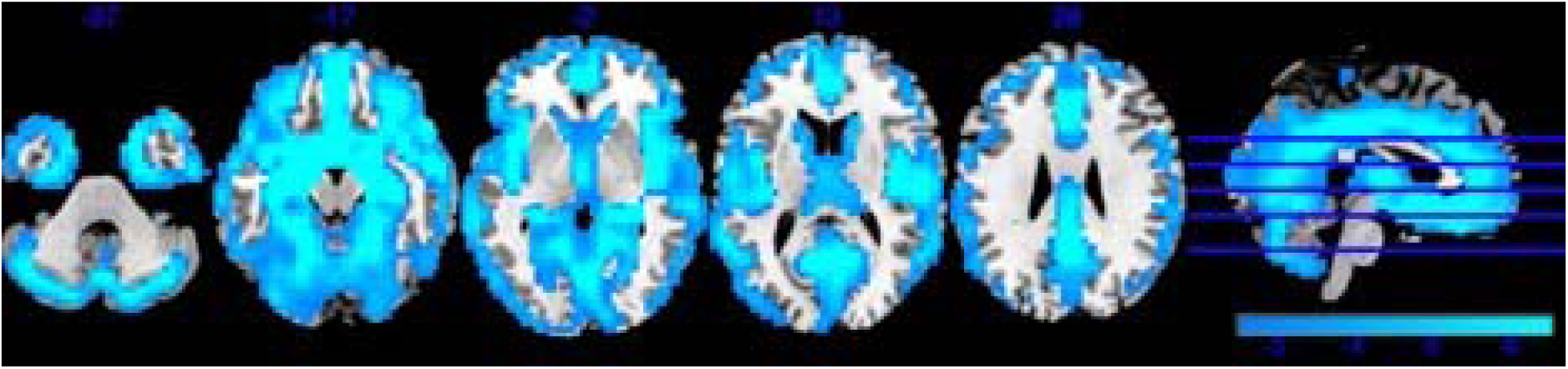
Gray Matter Alteration in Mild Cognitive Impairment

### Spectrum Results

Individuals classified as spectrum (N = 96) showed GM increases in the bilateral pallidum and putamen, and the left inferior occipital lobe. There were decreases in the GM of the bilateral insula, temporal poles, ventromedial prefrontal cortex and cingulum. See Figure 8 for more details.

**Figure 8.**
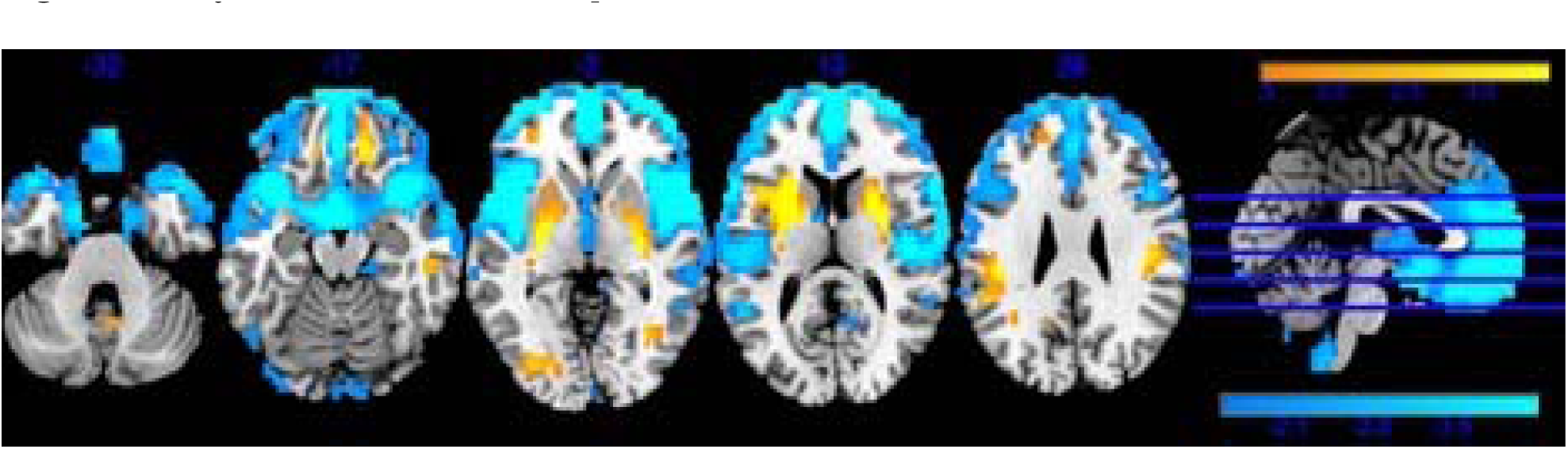
Gray Matter Alteration in Spectrum

### Unaffected Relatives

Individuals with first-degree relatives with psychosis (N = 147) had one of the only patterns of global GM increases across the cortex. Results show GM increases in the temporal poles, supplemental motor regions, cerebellum VI, precuneus and occipital lobes. There was also a small cluster of GM reductions in the right hippocampus. See Figure 9 for more details.

**Figure 9.**
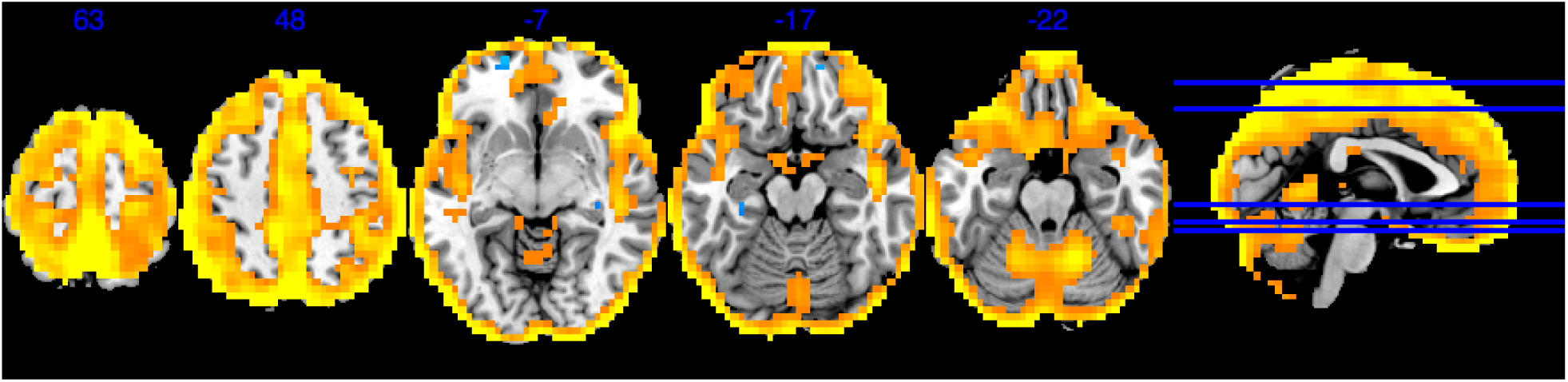
Gray Matter Alteration in Unaffected Relatives

### Spatial Correlations

The spatial correlations are displayed in a heat map distribution in Figure 10. There was an overall significant positive correlation between the groups at the whole-brain level; SZ and the spectrum group (*r* = .79), SZ and MDD (*r* = .72), and ASD and SZ (*r* = .69). As mentioned above, individuals with SZ had global reductions across the entire cortex. Individuals with MDD had gray matter reductions in the cingulum, bilateral insula, rectus, amygdala, and Heschl’s gyrus. SZ and MDD overlapped in the bilateral insula (peak: x = 37, y = 11, z = −12) and the medial prefrontal cortex. SZ and spectrum overlapped in a similar pattern, both groups had reduced GM concentration in the bilateral insula, ventromedial prefrontal cortex, and the cingulum. SZ and ASD overlapped in GM reductions in the olfactory bulb, parahippocampal gyrus, and to a lesser extent, the bilateral insula.

**Figure 10.**
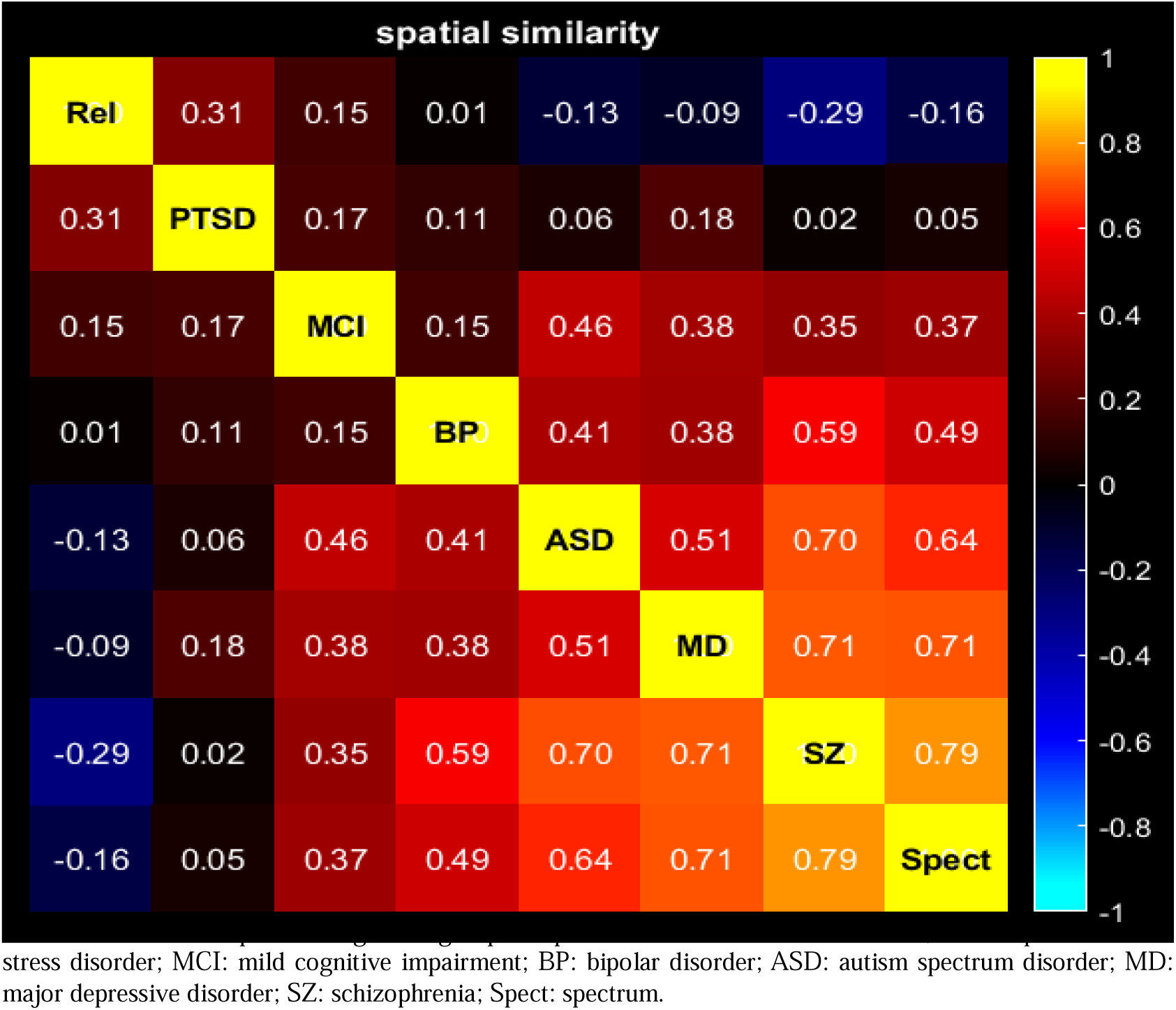
Heat Map of Spatial Correlations. Correlations for each pair of diagnostic group comparisons. Rel: Unaffected relatives, PTSD: post-traumatic stress disorder; MCI: mild cognitive impairment; BP: bipolar disorder; ASD: autism spectrum disorder; MD: major depressive disorder; SZ: schizophrenia; Spect: spectrum.

Unaffected relatives showed general increases in GM across the cortex and therefore, were anticorrelated with ASD (*r* = −.13), SZ (*r* = −.29), and to a lesser extent, MDD (*r* = −.09). Individuals with PTSD were largely uncorrelated with all other disorders even though there were decreases observed in the left insula and bilateral superior occipital gyrus. Individuals with PTSD were positively correlated with unaffected relatives (*r* = .31). Overlap between unaffected relatives and PTSD was largely seen in the superior parietal lobule and the motor region and the posterior insula.

### Post-hoc Analysis

We completed a subsequent post-hoc analysis on the spatial correlations with MCI individuals removed because of the potential for cognitive deficits to underlie psychiatric disorders (Figure 11). The directionality did not change for any correlation. In the post-hoc analyses, unaffected relatives were more anticorrelated with spectrum (*r* = −.24), SZ (*r* = −.37), MDD (*r* = −.16), and ASD (*r* = −.23).

**Figure 11.**
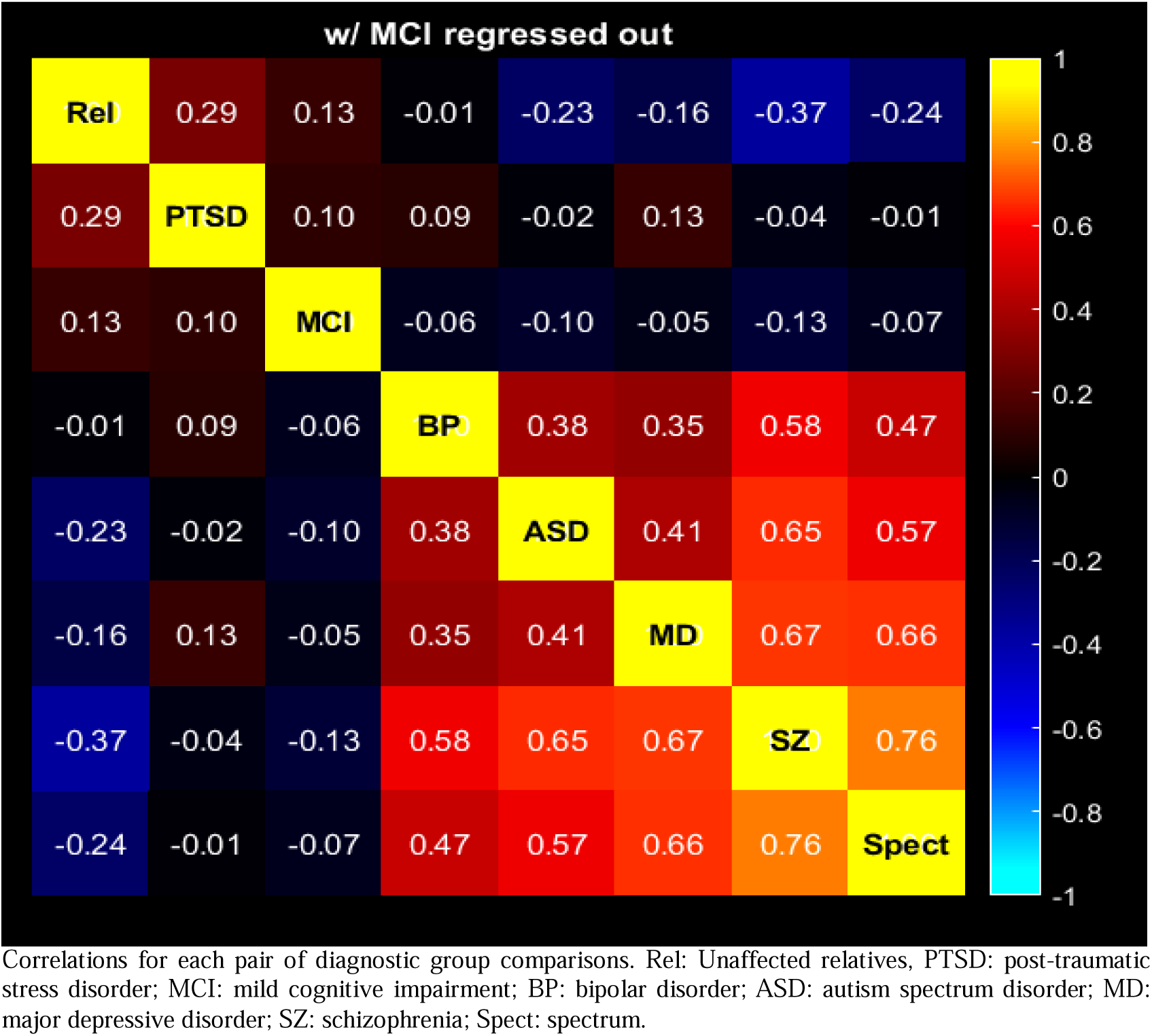
Heat Map of Spatial Correlations with MCI Correlated Out of Comparisons. Correlations for each pair of diagnostic group comparisons. Rel: Unaffected relatives, PTSD: post-traumatic stress disorder; MCI: mild cognitive impairment; BP: bipolar disorder; ASD: autism spectrum disorder; MD: major depressive disorder; SZ: schizophrenia; Spect: spectrum.

## Discussion

The goal of this paper was to examine the underlying neuroanatomic alterations across several diagnostic categories as well as their similarity to one another via the use of a decentralized analysis of large datasets present across multiple sites. Examination across multiple disorders allows for identification of possible inter-diagnostic similarities as well as a possibly protective finding by comparing healthy volunteers to all diagnostic groups. In the current large-scale cross-disorder study we identified both shared and disease-specific gray matter alterations in SZ, BP, MDD, ASD, MCI, and PTSD. In comparing healthy volunteers to each diagnostic category, we identified regions of significant gray matter reductions spanning the bilateral insula, hippocampus, and prefrontal cortex that showed significant differences when compared to healthy volunteers.

Individuals with SZ had the largest effect size of all diagnostic groups and overall global reductions throughout the cortex when compared to HCs. Our results largely support previous SZ findings of gray matter reductions in the bilateral insula, prefrontal cortex, cerebellum, and across the limbic system (Aine et al., 2017; Fornito et al., 2011; Gupta et al., 2015). Individuals with MDD had a similar GM pattern to SZ, with strong correlation between these spatial *p*-value maps. Previous studies have reported that initial, prodromal symptoms of SZ are low mood and anhedonia. Our findings support the notion that these disorders may have a similar neuroanatomical underpinning.

Individuals with PTSD did not have significant overlap in affected cortical regions with any other examined disorders. The alterations in PTSD appear unique and may indicate that PTSD is dissimilar to other psychiatric disorders in terms of biological basis. We found that the retrosplenial cortex was an area of reduced gray matter concentration in individuals with PTSD. The retrosplenial cortex has been identified as a region relating to multiple sensory functions including navigation (Alexander et al., 2023). The anatomical location also indicates that it is a bridge for regions involved in sensorimotor processing and spatial processing. A recent review categorized the main functions as perspective shifting across a multitude of spatial reference frames and prediction updating and generation from highly complex spatiotemporal contexts (Alexander et al., 2023). Given the unique nature of PTSD developing following a traumatic experience, the alterations we observed may be relating to a stress-response and not *just* a biological vulnerability to psychiatric disorder. In our study, there was an unexpected positive correlation between PTSD and unaffected relatives with significant overlap identified in GM increases in the superior parietal lobule. The superior parietal lobule has been identified as a hub for visual and sensory input which may relate to some of the symptoms of PTSD (e.g., hypervigilance) (Howard et al., 2000; Kunimatsu et al., 2020).

A recent study identified increases in the bilateral putamen of multiple psychiatric disorders when compared to healthy controls (Gong et al., 2019). Interestingly, this volumetric increase was also identified within their unaffected relatives (Gong et al., 2019). We also found a significant cluster in the left putamen in our group of unaffected relatives. The putamen is thought to serve as a center for the integration of high-level cognitive, motor, and limbic processes by receiving afferent inputs from other cortical regions and then transmitting them back to the neocortex through the thalamus (Bernácer et al., 2012). Given these functions, dysregulation of this region is thought to underlie the emergence of some of the more severe cognitive and clinical symptoms in psychiatric disease (Simpson et al., 2010). Previous studies have reported larger gray matter volume in the putamen of individuals with SZ, relative to healthy controls (McCarley et al., 1999; van Erp et al., 2014). Our results showed a significant increase in gray matter concentration in the putamen of unaffected relatives and to a much smaller extent in individuals with SZ, ASD, and spectrum.

### Significant Findings in the Insula

In our study, the insula was identified as an affected region in SZ, BP, MDD, ASD, PTSD, spectrum, and MCI. These findings are not surprising given the functionality of the subregions of the insula; the anterior and posterior insula (Centanni et al., 2021). The anterior lobe has largely been associated with executive control and sensory processing (Craig, 2009; Menon & Uddin, 2010; Molnar-Szakacs & Uddin, 2022). Dysfunction in this region may relate to altered orientation towards salient information, an often referenced explanation for psychosis as well as autism spectrum disorder. In functional studies, the anterior insula cortex was activated during errors in performance and error awareness (Harsay et al., 2012; Klein et al., 2007; Ullsperger et al., 2010). Specifically, the insula-cortico-thalamic circuit, including the dorsal and ventral areas of the anterior insula, is responsible for both error awareness and the processing of salience (Harsay et al., 2012). In addition to motivated behaviors, the insula is also considered significant for attention and emotional regulation. The anterior insula has a significant role in empathy and previous functional MRI studies have reported activation in this region in response to seeing other individuals in pain as well as to expressions of extreme emotions (e.g., fear and happiness) in others (Uddin et al., 2017). The dorsal mid-insula is also responsible for interoception, the detecting and regulating of the body’s internal state (Craig, 2002). Disruptions in interoceptive processing has previously been linked to multiple psychiatric disorders and a vulnerability to certain psychiatric symptoms (Craig, 2009; Nord et al., 2021; Petzschner et al., 2017; Quadt et al., 2018; Sheets et al., 2013). In SZ, previous research found reduced connectivity in the insula and that differences in those connectivity profiles were related to different symptom profiles (Tian et al., 2019). Taken together, our results and previous research indicate that the bilateral insula, with its numerous roles and connections to other vital subcortical regions, may have a pathological role in psychiatric disorders (Jiang et al., 2021; Meda et al., 2015; Namkung et al., 2017; Segall et al., 2009). Future research should explore the association of the insula and its related networks in not only SZ but other psychiatric disorders.

Other regions identified within this study as potential regions of biological vulnerability support the long list of previous literature examining structural alterations in psychiatric disorders. Even in disorders with significant phenotypic heterogeneity (e.g., SZ) we were able to identify consistent alterations within the rectus, which has previously been associated with a genetic risk for SZ, BP, and psychosis (Luna et al., 2022), and the cerebellum, with varying regions associated with cognitive losses (e.g., long-term and working memory) and psychosis (Clark et al., 2020; Moberget et al., 2018; Moberget & Ivry, 2019). Our findings within the hippocampus are also aligned with previous literature (Brosch et al., 2022; Lorenzetti et al., 2009; McCutcheon et al., 2023; Torres et al., 2016) that found reductions within MDD, BP, and SZ. Other cross-disorder studies found similar shared and disease-specific alterations in the hippocampus, amygdala, thalamus, accumbens of individuals with SZ, BP, and MDD (Okada et al., 2023). Future research is needed to examine these alterations in the whole-patient context to identify their relationships to cognitive and social functioning, course of illness, and medication.

## Limitations

There are a few limitations in this study. To start, the current study is cross-sectional in design, therefore, the potential changes in alterations over the course of disorder were not examined. Future studies should explore cross-disorder presentations using longitudinal approaches. Next, we did not examine additional social and environmental determinants such as duration of illness, medication, symptom profiles, race/ethnicity, or socio-economic status as these factors were not a focus of the current study. We acknowledge that these factors may have significant impacts on diagnostic presentations and gray matter alterations. Future research should examine these differences further and could also benefit from directly comparing disorders to other another (as opposed all disorders compared to healthy controls), a method we were not able to explore in the current study given insufficient power. We also did not match the groups for age or sex and acknowledge that there were significant variations in those demographics across each diagnostic group. However, our results still showed significant differences across diagnostic groups indicating valid representation of gray matter alterations across different disorders. The inclusion of age and sex on the regression models may have ameliorated the impact of these cohort difference to some extent, as our findings were still significant after accounting for these covariates.

## Conclusions

Through federated analyses within COINSTAC, we were able to identify gray matter patterns that were disorder specific as well as those that overlapped across different psychiatric disorders. Overall, the findings followed a pattern of gray matter reductions in all examined disorders. The reductions were largely seen in the bilateral insula, medial prefrontal cortex, parahipppocampal gyrus, and rectus. There were some noted increases in gray matter concentration in the regions of the putamen and pallidum. We identified consistent overlap in alterations within the insula across all disorders. Given the functionality of the insula, we hypothesize that this region may play a critical role in the vulnerability to psychiatric disorders. We acknowledge that there is likely not a *single* point of vulnerability or deficit within the cortex that will solely explain the presence of a specific psychiatric disorder, let alone all mental illnesses. However, transdiagnostic comparisons allow for greater understanding of the physiological and neuroanatomical bases of psychopathology and our results, which show an overlapping pattern within the insula across SZ, ASD, MDD, BP, PTSD, and MCI, which may point to a transdiagnostic locus of disruption. COINSTAC allowed for this federated approach and may be a useful tool for classification models or additional transdiagnostic analyses in the future.

## Data Availability

All data produced in the present study are available upon reasonable request to the authors.

