## Supplemental Material for "Cortical Similarities in Psychiatric and Mood Disorders Identified in Federated VBM Analysis via COINSTAC"

*Recruitment and Inclusion/ Exclusion criteria*

**ASRB.** Inclusion: All participants were fluent English speakers and aged 18-65 years old.

Exclusion: No history of an organic brain disorder, brain injury accompanied by > 24 h of amnesia, mental retardation defined as an IQ < 70, movement disorder, current substance dependence, or electro-convulsive therapy in the preceding 6 months. The control participants additionally had no personal history of psychotic disorder or family history of psychotic disorder in their first-degree biological relatives.

**COCORO.** Participants recruited from the Osaka site had no biological relations, and all of them were of Japanese descent. The subjects were excluded if they had neurological or medical conditions that could potentially affect the central nervous system, such as atypical headaches, head trauma with loss of consciousness, chronic lung disease, kidney disease, chronic hepatic disease, thyroid disease, active cancer, cerebrovascular disease, epilepsy, seizures, substance-related disorders, or mental retardation. Patients with schizophrenia, bipolar disorder, autism spectrum disorder, and major depressive disorder were recruited from the Osaka University Hospital. Each patient had been diagnosed by at least two trained psychiatrists according to the criteria from the diagnostic and statistical manual of mental disorders, fourth edition (DSM-IV) based on the structured clinical interview for DSM-IV (SCID). Controls were recruited through local advertisements at Osaka University. Healthy comparison subjects were evaluated using the non-patient version of the SCID (Gorgens, 2011) to exclude individuals who had current or past contact with psychiatric services or who had received psychiatric medications.

**Emory.** See (Fani et al., 2012) for recruitment and inclusion/exclusion criteria.

**IGP.** Inclusion: All participants were fluent English speakers and aged 18-65 years old

Exclusion: General exclusion criteria included an inability to communicate sufficiently in English, a current neurological disorder, a diagnosis of substance abuse or dependence in the past six months; and/or having been treated with electroconvulsive therapy in the previous six months.

**MAS.** See (Jiang et al., 2018) for recruitment and inclusion/exclusion criteria.

**OATS.** See (Sachdev et al., 2009) for recruitment and inclusion/exclusion criteria.

**Sydney.** Control (CON) participants were defined as those who did not have a first-degree relative with either BD I or II, recurrent major depressive disorder (MDD), schizoaffective disorder, recurrent substance abuse or any past psychiatric hospitalization. Additionally, they did not have a second-degree relative with a history of psychosis or who had been hospitalized for a mood disorder.  High-risk (HR) and CON participants with a lifetime or current presence of psychiatric symptoms (apart from the occurrence of BD) were not excluded from the study. This ecological approach has been used by similar studies of individuals at high genetic risk for BD to recruit both CON and HR cohorts (Nurnberger et al., 1994). BD participants had a BD I or II diagnosis. Recruited participants are involved in an ongoing longitudinal study with annual follow-up evaluations. HR participants were recruited from families who had either previously participated in a bipolar disorder (BD) pedigree molecular genetics study, from a specialized BD research clinic, or were otherwise recruited from clinicians, mental health consumer organizations and other forms of publicity. CON participants were recruited via print and electronic media as well as noticeboards in universities and local communities.

*Spectrum Diagnoses*

**ASRB.** All individuals labeled as spectrum had a psychotic disorder. There were eleven individuals with a diagnosis of psychotic disorder Not-Otherwise-Specified (NOS) and three with a diagnosis of depressive disorder with psychotic features.

**COCORO.** All individuals labeled as spectrum fell into one of the following diagnostic categories: ADHD, personality disorders, adjustment disorders, somatoform disorders, dissociative disorders, mood dysphoria, social anxiety, generalized anxiety, obsessive-compulsive disorder, learning disabilities, intellectual disabilities, borderline intelligence, eating disorders, falsehood disorders, organic mental disorders, substance-induced psychotic disorders.
